# Anticipating the curve: can online symptom-based data reflect COVID-19 case activity in Ontario, Canada?

**DOI:** 10.1101/2021.01.15.21249879

**Authors:** Arjuna S. Maharaj, Jennifer Parker, Jessica P. Hopkins, Effie Gournis, Isaac I. Bogoch, Benjamin Rader, Christina M. Astley, Noah Ivers, Jared B. Hawkins, Liza Lee, Ashleigh R. Tuite, David N. Fisman, John S. Brownstein, Lauren Lapointe-Shaw

## Abstract

**Background:** Limitations in laboratory diagnostic capacity and reporting delays have hampered efforts to mitigate and control the ongoing COVID-19 pandemic globally. Syndromic surveillance of COVID-19 is an important public health tool that can help detect outbreaks, mobilize a rapid response, and thereby reduce morbidity and mortality. The primary objective of this study was to determine whether syndromic surveillance through self-reported COVID-19 symptoms could be a timely proxy for laboratory-confirmed case trends in the Canadian province of Ontario.

**Methods:** We retrospectively analyzed self-reported symptoms data collected using an online tool – Outbreaks Near Me (ONM) – from April 20^th^ to Oct 11^th^, 2020 in Ontario, Canada. We estimated the correlation coefficient between the weekly proportion of respondents reporting a COVID-like illness (CLI) to both the weekly number of PCR-confirmed COVID-19 cases and the percent positivity in the same period for the same week and with a one-week lag.

**Results:** There were 314,686 responses from 188,783 unique respondents to the ONM symptom survey. Respondents were more likely to be female and be in the 40-59 age demographic compared to the Ontario general population. There was a strong positive correlation between the weekly number of reported cases in Ontario and the percent of respondents reporting CLI each week (r = 0.89, p <0.01) and with a one-week lag (r = 0.89, p <0.01).

**Interpretation:** We demonstrate a strong positive and significant correlation (r = 0.89, *p* <0.01) between percent of self-reported COVID-like illness and the subsequent week’s COVID-19 cases reported, highlighting that a rise in CLI may precede official statistics by at least 1 week. This demonstrates the utility of syndromic surveillance in predicting near-future disease activity. Digital surveillance systems are low-cost tools that may help measure the burden of COVID-19 in a community if there is under-detection of cases through conventional laboratory diagnostic testing. This additional information can be used to guide a healthcare response and policy decisions.

## INTRODUCTION

Severe acute respiratory syndrome coronavirus 2 (SARS-CoV-2) and the associated disease, coronavirus disease 2019 (COVID-19), have spread rapidly since first documented in China in late 2019^1^. Viral surveillance is an important public health tool that can help detect outbreaks, mobilize a rapid response and thereby reduce morbidity and mortality^2,3^. However, there are limitations to relying solely on laboratory testing for COVID-19 surveillance. At an individual-level, delays between symptom onset and testing, and between testing and COVID-19 test results mean that reported cases typically reflect disease activity from 1-2 weeks prior^4^. When case counts are high, testing restrictions may be implemented to preserve capacity. Typically these have included prioritizing those with the highest pre-test probability for a positive result (e.g., symptomatic and/or exposure to a confirmed case) or those at risk of severe illness ^5^. Changes in COVID-19 testing volumes over time make it difficult to interpret any trends in confirmed case counts. Surveys from the first wave of the pandemic estimated that only 2-9% of Canadians with symptoms consistent with COVID-19 were tested^6^. When viral transmission and new case counts are high, further delays in testing and results may reduce the reliability of confirmed case data for identifying key epidemiological events such as exponential growth or curve flattening. These limitations highlight the need for more timely, comparable, and comprehensive methods of population disease surveillance to inform public health measures.

Syndromic surveillance is a public health tool used extensively to identify the beginning of seasonal influenza outbreaks in the United States and Canada, and for the surveillance of other viral and bacterial diseases globally^7–10^. Participatory surveillance, a subtype of syndromic surveillance, allows individuals to self-report symptoms through phone or internet-based applications^11^. Where testing is incomplete, crowd-sourced symptom data for COVID-19 can be used as a proxy for confirmed case counts, help to estimate the true burden of disease, and forecast future epidemiological trends with strong spatial and temporal resolution^12–14^. There have not been any studies comparing trends in self-reported symptoms through participatory syndromic surveillance with laboratory-confirmed COVID-19 cases in Canada. The primary objective of this study was to determine whether self-reported COVID-19 symptoms could be used as a timely proxy for laboratory-confirmed case trends in the Canadian province of Ontario.

## METHODS

### Overview and Setting

We retrospectively analyzed self-reported participatory surveillance COVID-19 symptoms and test results, in addition to laboratory-confirmed COVID-19 case and testing data from Ontario, Canada. Ontario is Canada’s largest province, with approximately 14.5 million residents. The first case of COVID-19 in Ontario was reported on Jan. 25^th^, 2020, and community transmission was estimated to have started on March 17^th^, 2020. To date, the province has experienced two waves of COVID-19 cases with the first wave peaking in mid-April at weekly average of approximately 600 daily cases and an 8% test positivity rate. The second wave is ongoing at the time of writing this work. This study was approved by the Ethics Review Board of University Health Network and the University of Toronto and a waiver of informed consent was granted because the data were collected for public health surveillance purposes. All methods were performed in accordance with institutional guidelines and regulations.

### Data Sources and Study Population

The four data sources used for this study include: 1) participatory surveillance survey data from Outbreaks Near Me (ONM), 2) participatory surveillance survey data from FluWatchers, 3) regional COVID-19 case reports from the Ontario Case and Contact Management Plus (CCM Plus) and 4) regional laboratory testing data from the Ontario Laboratory Information System (OLIS). We created weekly tabulations of syndromic survey data, COVID-19 case counts and laboratory tests using the International Organization for Standardization (ISO) week date, a time standard that ordinally specifies each week of the year, starting on Monday and ending on Sunday^15^.

Outbreaks Near Me (outbreaksnearme.org), formerly COVID Near You, is a web-based participatory health surveillance tool created by infectious disease epidemiologists at Boston Children’s Hospital and launched in March 2020. This team also created Flu Near You (flunearyou.org), a similar tool for influenza symptoms, which has been validated against clinical data sources and applied to predict influenza trends^7–9^. Participants are asked to report on present symptoms, date of symptom onset, demographic information, healthcare encounters, testing, and results. Respondents reported symptoms on the ONM website and could opt to leave their cell phone number to receive SMS reminders to complete the survey again every three days after their initial submission. The mean Canada-wide weekly response rate to the ONM SMS survey prompts was 65.2% (SD: 14.5%). Symptoms of possible COVID-19 were defined using the CDC Surveillance Case Definition for COVID-19 from the National Notifiable Diseases Surveillance System (NNDSS). The COVID-like illness (CLI) metric approved on August 5^th^, 2020 ^16^ is defined as the presence of at least two of: fever (measured or subjective), chills, rigors, myalgia, headache, sore throat, or at least one of: cough, shortness of breath, difficulty breathing, new olfactory disorder, or new taste disorder. This case definition had a reported sensitivity of 97-98% and a specificity of 33-43% in adults for detecting a COVID-19 diagnosis^17^. We identified repeat responses by age/sex/phone number and included only one response per person-week, prioritizing a CLI positive response and, if none occurred, the first response in each week. We included responses with a self-reported postal code originating from Ontario, Canada, between April 20^th^, 2020 (week 17) and October 11^th^, 2020 (week 41).

FluWatchers is an internet-based participatory surveillance tool created by the Public Health Agency of Canada in November 2015 to track Influenza-like Illness (ILI). Defined as the presence of fever and cough, ILI has a reported sensitivity of 51-54% and specificity of 86-90% for a COVID-19 diagnosis in adults^17^. Participants can sign up to receive weekly email reminders to report symptoms through a link to an online platform. A total of 7,690 users reported symptoms at least once between April 20^th^ and October 11^th^, 2020 in Ontario, and among these users, the average weekly response rate between weeks 17 and 40 was 71% (range 60-88%).

CCM Plus data system has been implemented in Ontario to record COVID-19 case information. Each of Ontario’s 34 public health units is responsible for local COVID-19 case investigation and entry of case information into CCM Plus. Ontario’s case definition for a confirmed case of COVID-19 has evolved based on science and available testing methods, but generally requires a positive laboratory test using a validated nucleic acid amplification test, including real-time PCR and nucleic acid sequencing. We obtained confirmed COVID-19 case counts from the CCM Plus data system on October 13^th^, 2020 for the time period between January 25^th^ (Ontario’s first case) and October 11^th^, 2020. Extracted de-identified data included case reported date, accurate episode date (date of symptom onset, or if not present the date of specimen collection), age and gender. We used the accurate episode date to estimate the date of symptom onset. We extracted a separate dataset from Ontario Laboratory Information System of the total daily COVID-19 tests by age and gender, with data ranging from January 23^rd^, 2020 to October 11^th^, 2020. Weekly percent positivity in Ontario was calculated by dividing total positive cases reported each week by the total number of tests reported each week.

## Analysis

### Demographics

We compared ONM respondent characteristics to those of the general Ontario population, and those undergoing COVID-19 testing in Ontario. Provincial population estimates on July 1^st^, 2020, by age and sex, were obtained from Statistics Canada^18^. Testing for differences in proportions was done using Chi-square tests and Fisher exact tests (if small cells). The age distributions of those reporting CLI and positive COVID-19 cases were plotted by week. Plots were also constructed for the weekly proportion of all ONM respondents and those with CLI who reported being tested for COVID-19.

### Syndromic trends

We plotted the weekly proportion of ONM respondents with ILI and compared it to the weekly proportion of FluWatchers respondents with ILI. We also calculated the association between these two variables using Pearson’s correlation coefficient and a t-test was used to determine its statistical significance.

We compared both the weekly percent positivity in Ontario and the weekly number of new reported cases against the proportion of ONM respondents reporting CLI a) one week prior and b) the same week. We used both contemporaneous and one-week lagged indicators because of the potential for participatory surveillance to anticipate provincial COVID-19 case data, particularly in light of the known delays between symptom onset and positive case reporting. We also compared the proportion of ONM respondents reporting CLI based on the week of symptom onset to the number of cases in Ontario based on the accurate episode date (a proxy for symptom onset date). For each of these, we reported Pearson’s correlation coefficient, and a determined statistical significance using a t-test. Clopper-Pearson confidence intervals were calculated and plotted as error bars for all proportions. The data were analyzed using R version 4.0.1 in the RStudio software environment, version 1.1.463 (RStudio Inc., Boston, MA). All testing for differences was done at a two-tailed *p* <0.05 significance threshold.

## RESULTS

### Outbreaks Near Me Respondents, April 20 - Oct 11^th^, 2020

There were 314,686 responses from 188,783 unique respondents to the ONM survey between April 20^th^, 2020 and October 11^th^, 2020. The total number of responses per week ranged from 3,849 - 10,454 with a mean of 7,115 weekly responses. The median reported age of respondents was 47 years (IQR 36-58) and 61.6% (n=32,417) were female. There was a significantly greater proportion of unique ONM respondents who identified as female (n = 32,417; 61.6% female) compared to the proportion of all Ontarians who received a test (n = 828,752; 60.0% female, *p* < 0.01) and compared to the general Ontario population (n = 7,371,442; 50.6% female, *p* < 0.01) (Table 1). The proportion of ONM respondents aged 40-59 years (n=23,432; 44.6%) was also significantly higher than that of the tested population (n = 410,229; 29.7%, *p* < 0.01) and the Ontario population overall (n = 3,915,662; 26.9%, *p* < 0.01). There was also a significantly smaller portion of respondents who were <19 years old in ONM (n = 1,816; 3.5%) compared to those who received a test (n = 9,4120; 6.8%, *p* < 0.01) and the Ontario general population (n = 3,141,693; 21.6%, *p* < 0.01). The age distribution of ONM respondents did not change over time. The <19 years age demographic consistently made up the lowest proportion of respondents, while the 40-59 age demographic was consistently the most likely to respond each week (Figure 1).

**Table 1.**
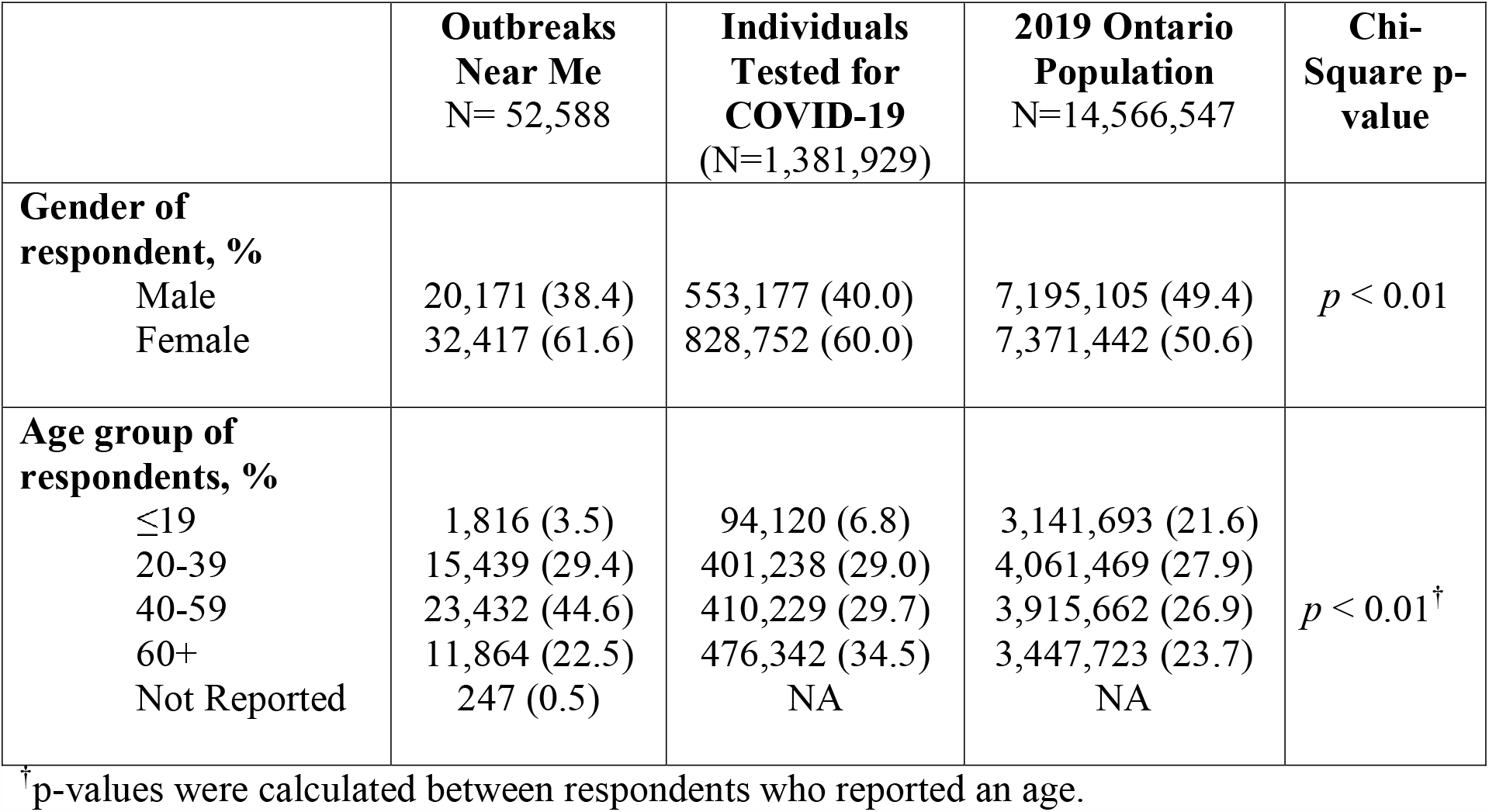
Self-reported characteristics of respondents in data sources compared to the Ontario Population

|  | **Outbreaks**<br>**Near Me**<br>N= 52,588 | **Individuals**<br>**Tested for**<br>**COVID-19**<br>(N=1,381,929) | **2019 Ontario**<br>**Population**<br>N=14,566,547 | **Chi-**<br>**Square p-**<br>**value** |
| --- | --- | --- | --- | --- |
| **Gender of**<br>**respondent, %** |  |  |  |  |
| Male | 20,171 (38.4) | 553,177 (40.0) | 7,195,105 (49.4) | $p < 0.01$ |
| Female | 32,417 (61.6) | 828,752 (60.0) | 7,371,442 (50.6) |  |
| **Age group of**<br>**respondents, %** |  |  |  |  |
| $\leq 19$ | 1,816 (3.5) | 94,120 (6.8) | 3,141,693 (21.6) | $p < 0.01^{\dagger}$ |
| 20-39 | 15,439 (29.4) | 401,238 (29.0) | 4,061,469 (27.9) |  |
| 40-59 | 23,432 (44.6) | 410,229 (29.7) | 3,915,662 (26.9) |  |
| 60+ | 11,864 (22.5) | 476,342 (34.5) | 3,447,723 (23.7) |  |
| Not Reported | 247 (0.5) | NA | NA |  |

<sup>†</sup> p-values were calculated between respondents who reported an age.

**Figure 1.**
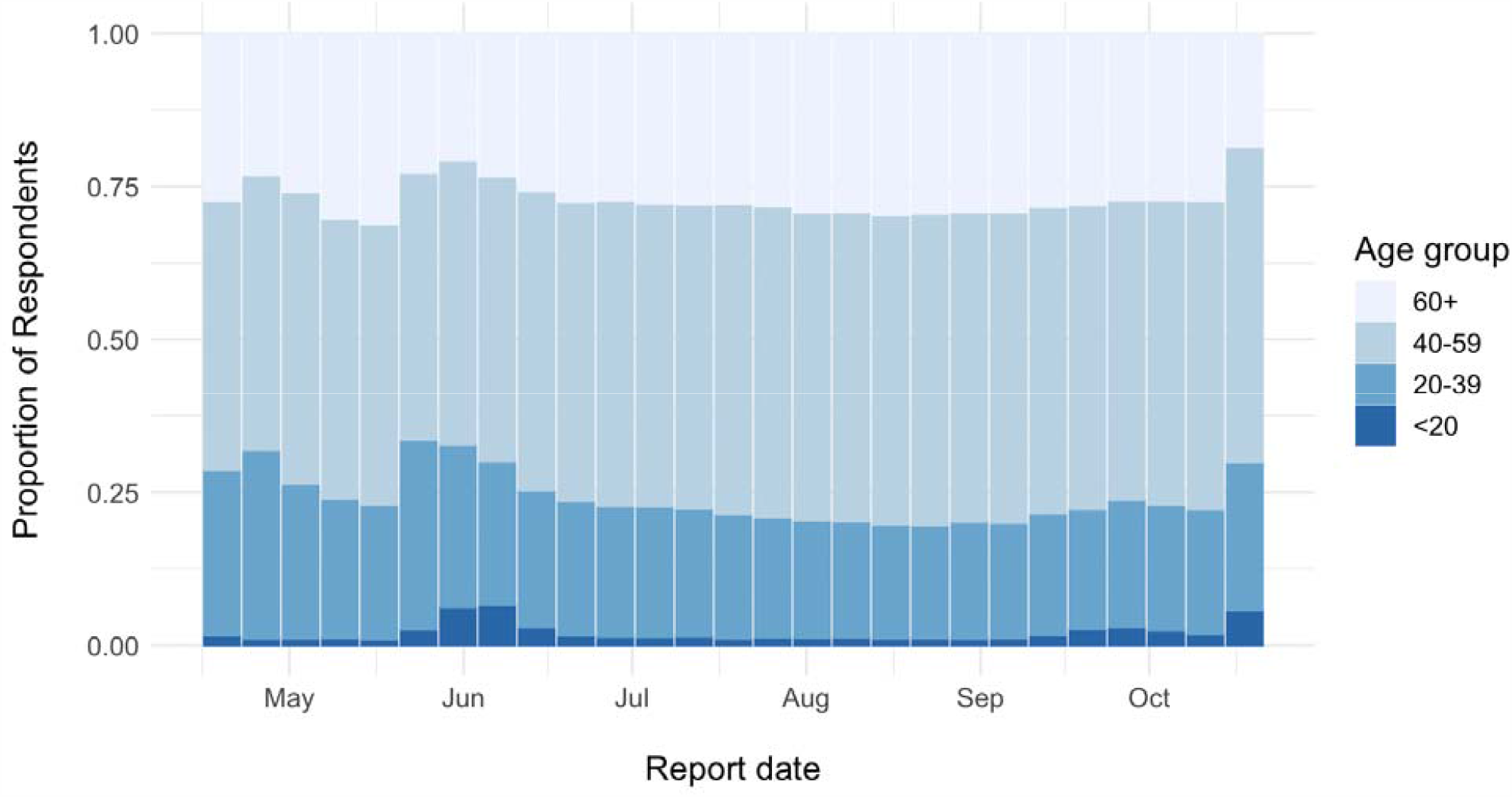
Age group of ONM respondents for ISO weeks 17 - 41

### Outbreaks Near Me Symptom Reporting

Overall, CLI was reported in 1.39% (n = 4,374) of responses, while 1.74% (n = 5,474) of all responses reported at least one symptom. The most commonly reported CLI symptom was fatigue (n = 1,382; 0.78%) and the least reported CLI symptom was loss of smell or taste (n = 364; 0.11%) (See Appendix Figure 1).

### Demographic trends of those reporting a COVID-like Illness to positive COVID-19 cases

The age groups of those reporting CLI changed over time. In the first five weeks of reporting (mid-April to mid-May) the 40-59 age group was the most likely to report CLI. Subsequently, the 20-39 year age group has consistently been the most likely to report CLI. Those in the <20 years age group showed an increasing trend in CLI reporting since the beginning of survey collection (Figure 2). Similarly, provincial COVID-19 case trends also exhibited an increase in the proportion of individuals <20 and 20-30 years of age from late April to September (Figure 3).

**Figure 2.**
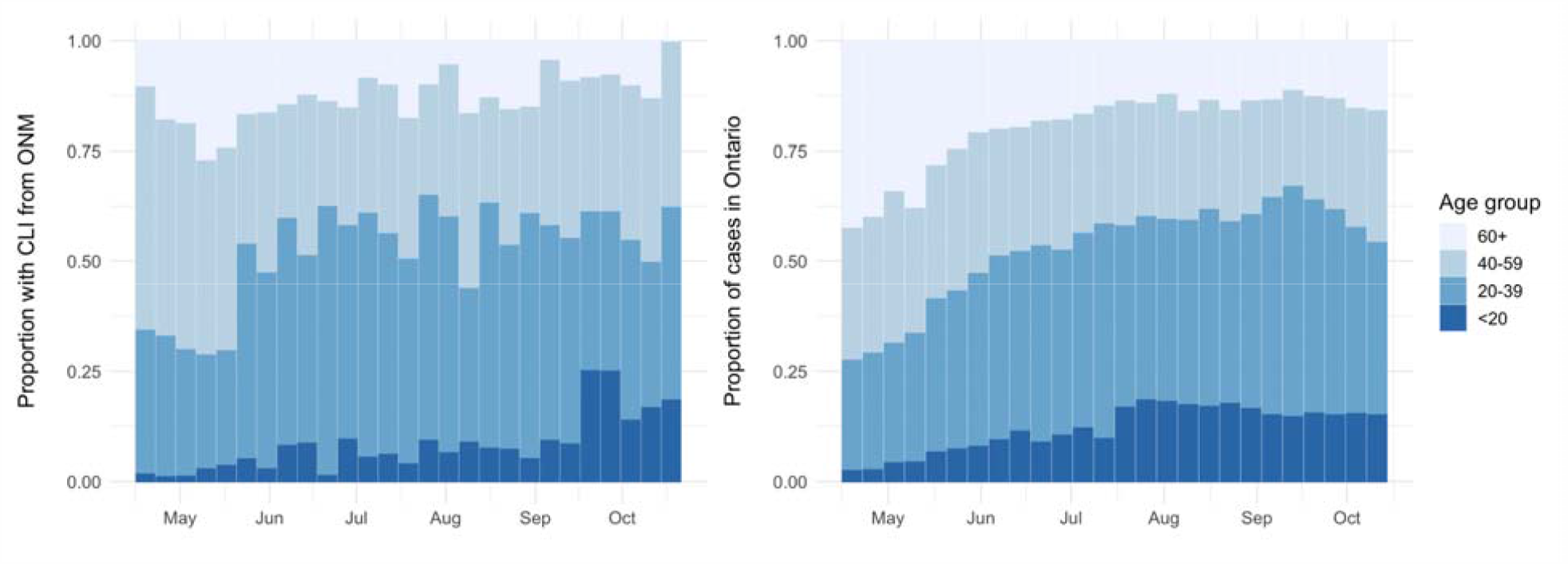
Reported age of those with CLI from ONM (left) and age of reported COVID-19 cases in Ontario (right)

**Figure 3.**
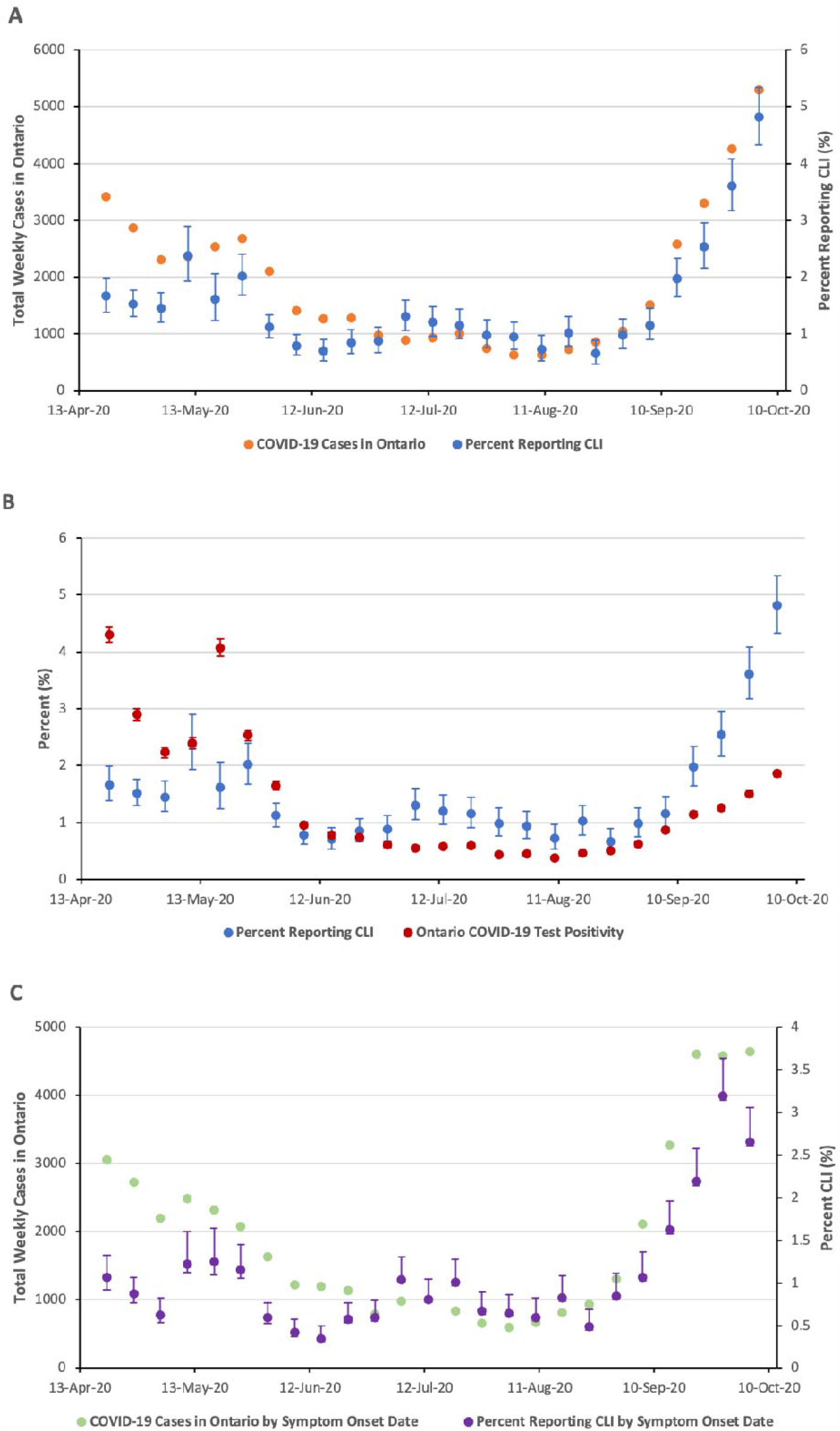
(A) Percent CLI and new COVID-19 Cases in Ontario each week. (B) Percent CLI and percent positivity for SARS-CoV2 in Ontario each week. (C) Weekly percent CLI and number o new COVID-19 cases based on the estimated date of symptom onset.

### Outbreaks Near Me Self-Reporting of Testing

Testing for COVID-19 was self-reported in 341 (0.2%) ONM survey responses with the weekly proportion self-reporting testing ranged from a high of 0.66% (n = 27) in ISO week 20 to a low of 0.04% (n = 3) in ISO week 33 (Appendix Figure 2). This reported rate of testing was similar to that of the Ontario population which ranged from 0.04 – 0.28% of the population tested each week. Among those who met the CLI symptom criteria (1592 responses), 16.3% (n = 260) reported receiving a COVID-19 test. Among those with CLI, the weekly proportion self-reporting testing ranged from a high of 28.9% (n = 20) in week 20 to a low of with 7.3% (n = 3) in week 36 (Appendix Figure 3).

### Correlation Between Survey and SARS-CoV-2 Data

#### Same Week

Similar trends were seen in the proportion of individuals reporting CLI and both the number of weekly new COVID-19 cases in Ontario (Figure 3A) and the COVID-19 test percent positivity in Ontario (Figure 3B). There was a weak positive correlation between the test percent positivity in Ontario and the proportion of respondents reporting CLI (r = 0.35, 95% CI [-0.05, 0.65], *p* = 0.08) and a strong positive correlation between the weekly number of reported cases in Ontario and the percent of respondents reporting CLI each week (r = 0.89, 95% CI [0.77, 0.95], *p* <0.01).

#### One-week lag

There was a strong positive correlation between the proportion of respondents reporting CLI and the number of new reported cases in Ontario the following week (r = 0.89, 95% CI [0.75, 0.95], *p* <0.01), and a moderate positive correlation between the proportion with CLI and the COVID-19 test percent positivity in Ontario the following week (r = 0.59, 95% CI [0.24, 0.80], *p* <0.01).

#### Symptom Onset

When used the estimated symptom onset date from both data sources, there was a strong positive correlation between the weekly proportion of respondents reporting CLI and the weekly number of new cases in Ontario (r = 0.87, 95% CI [0.72, 0.93], *p* <0.01) (Figure 3C).

### Correlation with Flu Watchers Data

The proportion of ONM respondents reporting ILI (fever and cough) each week ranged from a high of 0.21% (n=13) in week 39 (Sept. 21^st^ – 27^th^) to a low of 0% (n=0) in week 29 (July 13^th^ – July 19^th^. The proportion of respondents reporting ILI from ONM and from FluWatchers had similar ranges and trends over time (See Appendix Figure 4). There was a strong positive correlation in the weekly percentage of respondents reporting ILI between the ONM and FluWatchers survey (r = 0.73, 95% CI [0.61 – 0.85] *p* < 0.01).

## DISCUSSION

We found that there was a strong positive and significant correlation between self-reported COVID-like illness (CLI) and the subsequent week’s number of new COVID-19 reported cases, highlighting that the rise in CLI may precede official statistics by at least one week. This demonstrates the utility of syndromic surveillance in predicting near-future disease activity. Furthermore, ONM trends in fever and cough correlated with FluWatchers, another Canadian participatory surveillance system. These findings suggest there is the potential for participatory surveillance data to be useful for understanding and even forecasting COVID-19 trends in Ontario.

These findings build on previous reports of digital crowd-sourced surveillance techniques reported in the literature. Yoneoka et al. and Nomura et al. reported analyses of syndromic data collected through a large-scale (over 350,000 participants) digital surveillance system in Tokyo, Japan for one week. They observed a strong spatial correlation between symptoms and confirmed COVID-19 cases in Tokyo, Japan^12–14^. Here we show similar findings of self-reported symptoms tracking with COVID-19 confirmed cases for long-term data collected over several months in an entire Canadian province.

When comparing demographic characteristics between survey respondents and those who received COVID-19 testing, we found significant differences in age and gender. ONM respondents were more likely to be female and aged 40 – 59 years than those being tested for SARS-CoV2 in Ontario. Others have similarly reported that middle-aged females were the group most engaged with influenza participatory surveillance tools^7^. The ability of this survey to collect symptom data from groups other than those being tested allows for more comprehensive population coverage. Early limitations on testing to only the most severe cases caused uncertainties in true reported case counts^6^. Eligibility criteria for testing have changed over the course of the pandemic - most recently, testing was again restricted to those with symptoms, and only available by pre-scheduled appointment, effectively restricting the number of tests available^19^. Survey data that differs in demographic characteristics from those that received a test can provide valuable insights and fill knowledge gaps about groups not receiving testing. The addition of symptom surveillance information ensures that the most representative and timely data is used to inform policy decisions.

Participatory surveillance data also demonstrated that an increasing proportion of those reporting CLI, and of those testing positive for SARS-COV-2, were ages <20 and 20-39 years. In April those <40 years made up ∼25% of positive COVID-19 tests. As of October 2020, approximately 50% of positive COVID-19 tests in Ontario were individuals <40 years old. Similarly, approximately 50% of those with a COVID-like illness were <40 years of age at the end of our study observation period. These findings demonstrate the utility of ONM to detect the evolving demographic distribution of COVID-19.

We found that 0.12% – 0.66% of ONM respondents reported being tested for COVID-19. This was similar to the weekly Ontario population testing rate which ranged from 0.04% - 0.28% of the population. ONM respondents consistently reported a higher rate of COVID-19 testing than the Ontario general population. This may relate to ONM respondents taking a more active role in their health. We also found that a large proportion of respondents with COVID-like illness reported not being tested – with a decreasing trend since week 25. Among those who reported CLI, 16.3% reported being tested in the last 2 weeks, which dropped to 7.3% in week 37. In the first wave of the pandemic, we estimated that only 2-9% of those with symptoms had been tested^20^. Given growing awareness of the transmissibility and health impacts of COVID-19, one can only speculate as to why the testing rate among symptomatic people might be falling in the second wave. It is possible that due to a lack of paid sick leave, individuals may be less inclined to undergo testing due to the implications of a positive test on the inability to engage in employment.

There are several limitations to this study. Rapidly rising COVID-19 case counts are often broadcasted in the media and this can influence both healthcare-seeking behaviour and symptom self-awareness^21^. In the United Kingdom, significant national and local media coverage of COVID-19 cases resulted in observed increases in syndromic signals (both nationally and locally), particularly those associated with the symptoms of COVID-19 reported in the news media^21^. However, some participatory surveillance systems are more sensitive to the impact of media reporting than others – hence, a multi-source surveillance approach can be beneficial to reliably track population symptomatology. Next, the ONM participatory surveillance method relies on access to the internet, which may exclude individuals who are underhoused or experiencing homelessness, those with poor internet or computer access, or limited English literacy. These characteristics are more common among the racialized and marginalized groups who are disproportionately affected by COVID-19^22^. In addition, an estimated 17.9 – 30.8% of COVID-19 infected individuals are asymptomatic or minimally symptomatic^23,24^. The ability of syndromic surveillance methods to accurately predict case numbers will be confounded by changes in the rate of asymptomatic testing. This is because asymptomatic cases contribute to case counts but cannot be detected through syndromic surveillance methods. This study was also limited to Ontario. The generalizability of findings will depend on local uptake of syndromic surveillance tools, as well as on access to timely testing. In locations where testing is less accessible or characterized by long delays for test results, syndromic surveillance data may anticipate testing data by 1-2 weeks. Where testing is broadly accessible, with short delays, we expect present week symptom and case data to be more tightly correlated.

There have been few previous reports of an association between participatory surveillance signals and confirmed case counts for COVID-19^13,14^. This is the first such study to be reported in Canada. We found that online volunteer symptom surveys signalled subsequent increases in COVID-19 case counts in Ontario, Canada. Digital surveillance systems such as ONM are low-cost tools that may be helpful in determining the burden of COVID-19 in a community, particularly if there is under-detection of cases through conventional laboratory diagnostic testing. This additional information can be used as a cost-effective resource to guide a healthcare response and guide policy decision-making.

## Data Availability

Data will be made available upon request to the corresponding author

## Author Contributions

LLS, JSB, IIB, DNF, and ART led the conception and design of the work. BMR, ASM, and JP performed data extraction and analysis. LLS and ASM drafted the manuscript. ASM and ART prepared Figures and Tables. CMA, JP, JH, EG, LL, LLS, BMR, JBH, DNF, ART, IIB, JSB and NMI interpreted the data and provided critical review of the manuscript. All authors reviewed and approved the final manuscript.

## Acknowledgements

This study is funded by the UofT COVID action initiative (LLS). Outbreaks Near Me is funded by the Center for Disease Control and Prevention, Ending Pandemics and Flu Lab. FluWatchers is funded by the Public Health Agency of Canada.

## Appendix

**Appendix Figure 1:**
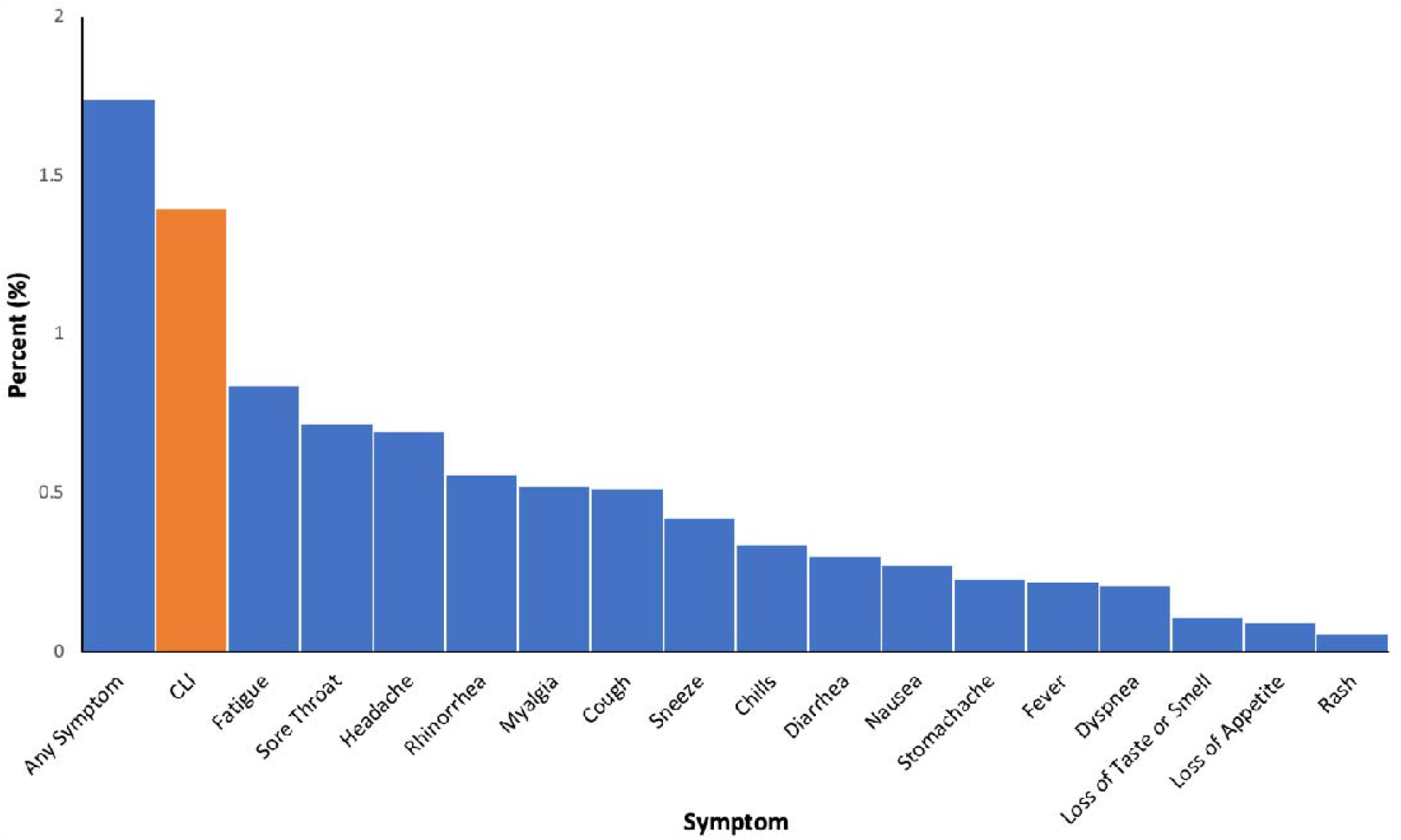
Percent of respondents reporting each symptom from the Outbreaks Near Me survey

**Appendix Figure 2.**
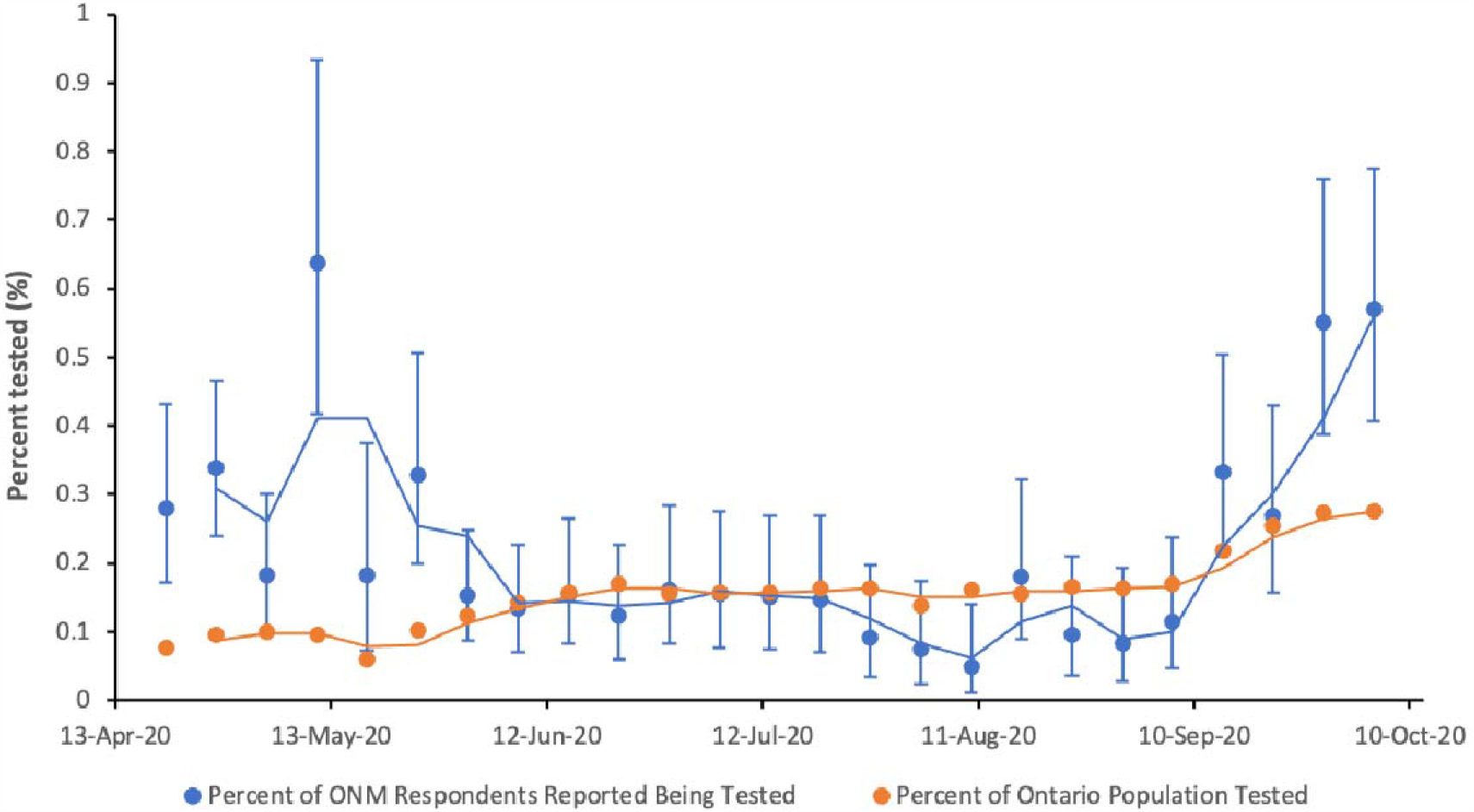
Percent of ONM responses reporting being tested, compared to the percent of the Ontario population tested for COVID-19, by week. Line indicates the two-week moving average.

**Appendix Figure 3.**
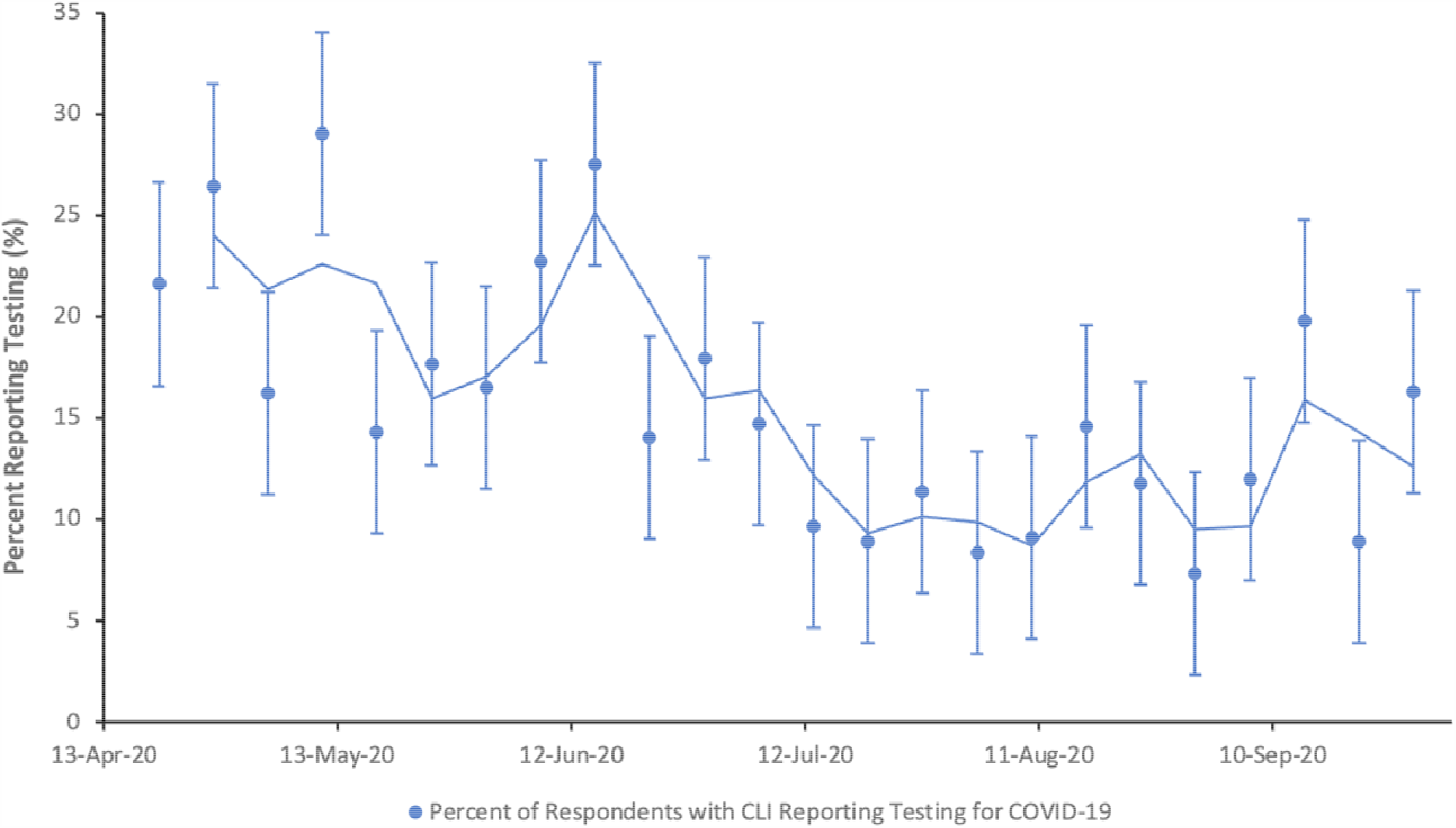
Percent of CLI-positive respondents reporting COVID-19 testing, by week. Line indicates the two-week moving average.

**Appendix Figure 4.**
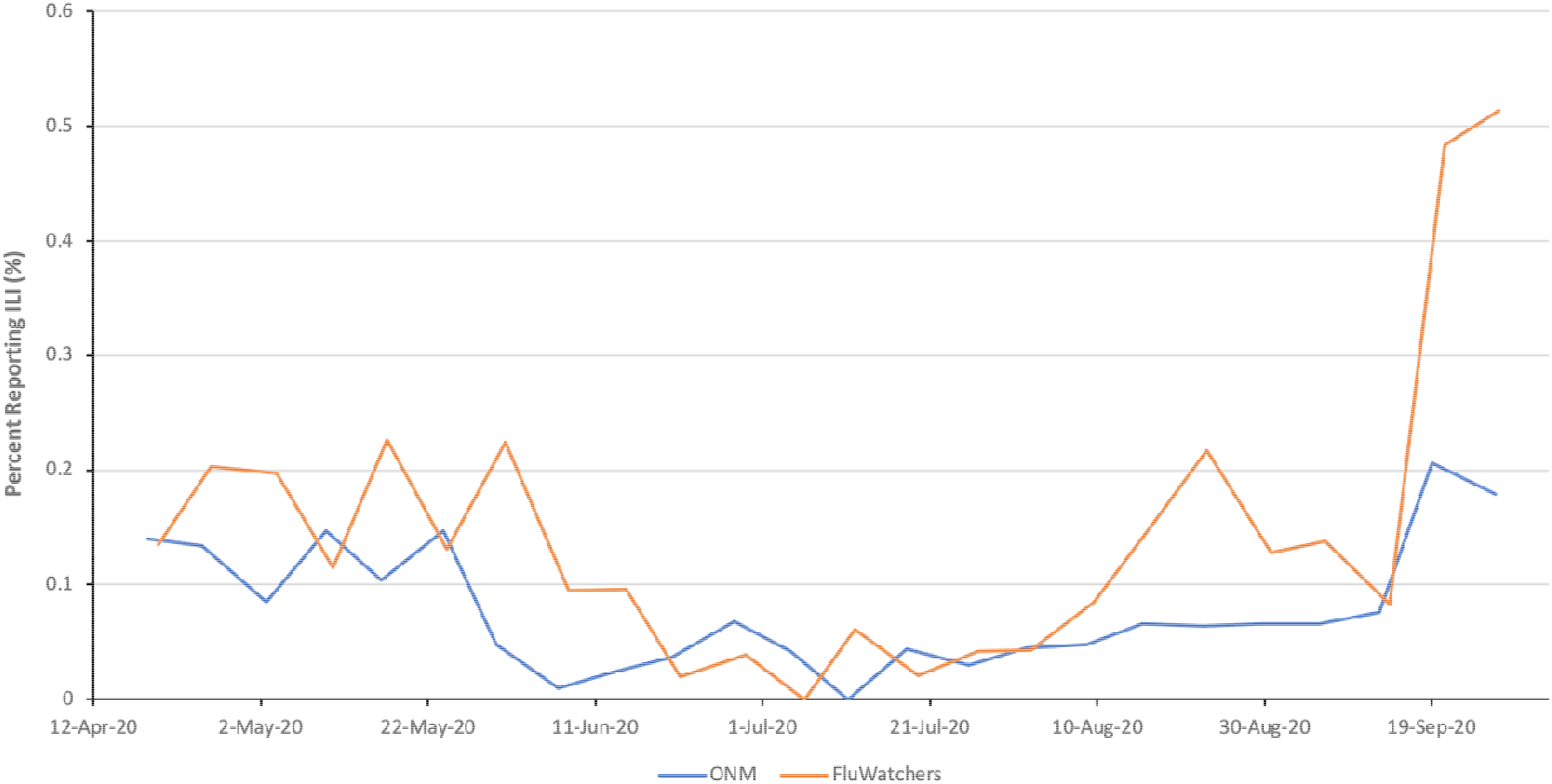
Proportion of individuals reporting ILI (both fever and cough) from Outbreaks Near Me and FluWatchers

